# Unequal Benefits: The Effects of Health Insurance Integration on Consumption Inequality in Rural China

**DOI:** 10.1101/2024.07.02.24309862

**Authors:** Linlin Han, Feng Yang, Jinxiang Yu

**Author notes:** **Correspondence:** Jinxiang Yu, Ph.D. School of Economics and Management, Nanjing Agricultural University, No.1 Weigang, Nanjing, Jiangsu Province,China. **CRediT authorship contribution statement** **Linlin Han**: Writing-review & editing, Writing-original draft, Supervision, Methodology, Formal analysis, Conceptualization. **Feng Yang**: Writing-review & editing, Resources, Funding acquisition, Conceptualization. **Jinxiang Yu**: Writing-review & editing, Project administration, Investigation, Formal analysis, Data curation.

## Abstract

Releasing the consumption potential of rural residents and narrowing the consumption gap is crucial for expanding domestic demand and enhancing social equity. This study leverages data from the China Family Panel Studies (CFPS) spanning the years 2012 to 2018 to analyze the impact of the rural-urban health insurance integration policy on consumption inequality in rural areas and its underlying mechanisms. Employing a staggered difference-in-differences (DID) approach, the analysis reveals that the policy significantly raises consumption levels among middle and high-income groups while concurrently reducing expenditures for the lowest-income bracket, thereby exacerbating consumption inequality. Heterogeneity analysis indicates that the impact of rural-urban health insurance integration on rural consumption inequality is manifested in both consumption structure and life-cycle effects, with the most significant disparities observed in subsistence and enjoyment consumption, particularly among middle-aged and older age groups. Mechanism analysis identifies increased utilization of medical services, the release of precautionary savings among middle and high-income cohorts, and variations in health insurance funding modalities as key drivers of the widening consumption inequality gap. The study concludes with recommendations to progressively advance the establishment of parity in rural-urban integrated health insurance and to prioritize policy support for vulnerable groups, especially the elderly and impoverished households.

## 1. Introduction

As China has become the largest developing country and its economy has achieved significant milestones, the aspiration for common prosperity amidst high-quality development has emerged. However, the country currently grapples with pronounced inequalities. The persistent expansion of disparities in resident income, wealth, and consumption, alongside the limited redistributive role of the social security system, has been a longstanding concern (Li and Zhu, 2018). These inequalities are evident not only in the differential access to welfare and public services across regions and between urban and rural areas but also in the social stratification within the same group regarding individual living welfare conditions. Consumption, a vital component of welfare, provides a comprehensive reflection of individual living standards. Specifically, consumption inequality can directly and vividly reveal the living welfare status of individuals. Compared to the higher-income groups with better health, the lower-income groups with poorer health face a distinct disadvantage in consumption expenditure. This individual-level consumption gap continues to damage the health capital of disadvantaged groups through the relative deprivation of their physiological health and the perception of psychological differences, leading to a range of diseases and long-term deprivation of their labor capacity. This exacerbates the existing consumption inequality and perpetuates it across generations(Deng and Yang, 2019). This situation further highlights the issue of wealth disparity and necessitates a focus on enhancing the consumption level of lower-income groups to effectively alleviate the consumption gap between different income levels, thereby mitigating social conflicts.

Equity and fairness are primary policy objectives in the healthcare domain. Despite being established since the 1990s, China’s medical insurance systems, including the Urban Employee Basic Medical Insurance, the New Rural Cooperative Medical Scheme for rural residents, and the Urban Residents Basic Medical Insurance, have had limited success in enhancing healthcare equity. In recent years, the Chinese government has pursued the integration of the New Rural Cooperative Medical Scheme with the Urban Residents Basic Medical Insurance into a singular framework known as the Urban-Rural Residents Medical Insurance (URMI). Data as of the year’s end in 2023 indicate that the URMI encompassed a population of 983.28 million, constituting roughly 69.84% of the country’s total populace. Consequently, URMI has emerged as the predominant medical insurance scheme in China in terms of the number of enrollees. The primary aim of this integration is to enhance the medical insurance entitlements for non-employed individuals in urban and rural areas, thereby promoting parity in medical insurance coverage between these two demographic groups. In accordance with the policy objectives, the integration of medical insurance should ideally lead to the dispersion of health risks among groups with varying levels of health and financial status. This is to be achieved through the refinement of income redistribution mechanisms, with the ultimate aim of reducing the socioeconomic disparities that can arise due to health-related issues. Nevertheless, the medical insurance system in China faces challenges due to limited economic capacity to provide a high level of equitable coverage. Consequently, the phenomenon of “reverse compensation,” where middle- and high-income groups disproportionately benefit from health services utilization compared to low-income groups, persists and remains a significant issue (Fan et al., 2021; Jin et al., 2020). However, previous studies have not yet assessed the policy effects of urban-rural health insurance integration from the perspective of consumption inequality. In addition, during the process of integrating urban and rural medical insurance, some regions have experienced issues such as delayed policy rollout, rapid increases in individual payment standards, and significant differences in existing systems, which hinder the income redistribution regulatory role of the integrated URMI (Gao and Wang, 2017). In light of this, this paper attempts to theoretically analyze and empirically test potential problems in the process of URMI integration reform and its impact on consumption inequality and its mechanisms, providing useful references for the improvement of the design of the medical insurance system.

Based on the perspective of consumption inequality, this paper explores the policy effectiveness of urban and rural medical insurance integration in rural areas. Initially, the characteristic fact of consumption relative deprivation within rural households is measured using the Kakwani Relative Deprivation Index, based on data from four phases of the CFPS. Subsequently, the impact of urban and rural medical insurance coordination on consumption inequality is empirically examined using the staggered DID model, with heterogeneity analysis across consumption structures and life-cycle dimensions. Finally, this paper assesses the mechanisms that may exacerbate consumption inequality in rural areas, including the utilization of health services, precautionary savings, and health insurance financing models.

The potential contributions of this study are as follows: First, it pioneers the evaluation of benefit equity within the healthcare system from a consumption perspective. Integrating consumption relative deprivation into the analytical framework leads the way in healthcare benefit equity, which traditionally focuses more on health-related inequalities. Second, it broadens the scope of consumption inequality research. While existing literature predominantly examines consumption gaps between urban and rural areas using macro indicators, this paper constructs relative deprivation indicators to assess consumption disparities at the individual level, providing a nuanced examination of consumption differences within rural areas. Third, it uses a precise method of causal identification. Considering the exogenous and progressive implementation characteristics of the urban and rural medical insurance coordination reform, the article constructs a staggered DID method with area and time fixed effects. Furthermore, this paper uses multiple scientific robustness tests to verify the causal relationship between urban and rural medical insurance coordination and consumption relative deprivation.

## 2. Background and literature review

### 2.1 Background

China has historically grappled with disparities in the distribution of medical insurance benefits. In response, several provinces, including Qinghai, Chongqing, Tianjin, Ningxia, and Guangdong, pioneered the integration of the Urban Residents Medical Insurance and the New Rural Cooperative Medical Scheme into a unified system post-2008. This initiative gained momentum in 2016 following the national government’s clear directives on medical insurance integration. The resulting integrated URMI has demonstrated significant enhancements in terms of reimbursement rates, designated medical facilities, pharmaceutical catalogs, and coverage for major illnesses, surpassing the previous standards. The guiding principles of the integration—prioritizing higher reimbursement rates, broader drug coverage, and a greater number of designated hospitals—have contributed to a reduction in the gap in medical insurance entitlements between urban and rural populations.

From a financial perspective, the funding models for integrated URMI across regions can be broadly classified into two categories: a uniform payment system and a tiered payment system (He and Shen, 2021). The uniform system requires uniform premiums and benefits for all insured individuals, whereas the tiered system permits adult enrollees to select from multiple premium levels, with benefits directly proportional to the contributions made. The majority of cities have established two premium levels based on the previous standards of Urban Residents Medical Insurance and the New Rural Cooperative Medical Scheme. A minority of regions have introduced three-tier or more complex systems, which either include an intermediate tier or cater to families with serious illnesses or higher economic status, aligning with the principles of the tiered system. The overarching aim of the URMI integration is to progressively improve medical insurance benefits for residents, narrow the gap in medical security coverage, and develop a more equitable and rational medical security system. The effectiveness of this policy in achieving equity has been a focal point of interest for both the state and the populace. However, given the recent completion of the integration process, research into the policy’s impact on equity remains in its infancy and requires further in-depth exploration.

### 2.2 Literature review

#### The trends and measurement of consumption inequality

Examining the trend of consumption inequality revealed a general increase over the past five decades across many countries(Attanasio and Pistaferri, 2014; Krueger and Perri, 2006). Cai et al. (2010) and Li et al. (2023) explored consumption inequality trends in China, revealing a pattern of initial rise followed by a decline. Measurement methods for consumption inequality can be categorized into group-level and individual-level indicators. Common group indicators include the Gini coefficient(Gayán-Navarro and Sanso-Navarro, 2024), the Atkinson index(Fischer and Lundtofte, 2020), the generalized entropy index(Dufour et al., 2024), and the interquartile ratio(Girsberger et al., 2020), which provide insights into societal or group-level consumption disparities. However, recent studies have increasingly focused on individual-level measurements to address consumers’ specific concerns about the relative deprivation they face. Individual-level indicators like the Yitzhaki index, the Podder index, and notably, the Kakwani index have gained prominence due to their dimensionless, regularized properties, with the mean being the Gini coefficient(Ledić et al., 2023; Pak, 2023; Deng and Yang, 2019). The Kakwani index calculates the consumption gap between an individual and all others in their reference group with higher consumption levels, offering insights into inequality induced by upward social comparisons within reference groups.

#### The causes of consumption inequality

From the perspective of household heterogeneity characteristics, income inequality emerged as a pivotal determinant of consumption inequality(Krueger and Perri, 2006). Increased property values played a role in mitigating consumption inequality among the youth, attributed to the wealth effect and the housing slave effect(Zhang et al., 2022). Financial literacy, by facilitating credit smoothing, asset appreciation, and insurance protection, reduced relative deprivation in rural households (French and McKillop, 2016). From the perspective of the external environment and institutions, factors such as state-owned multinational enterprises, urbanization, and globalization exerted influence on consumption inequality(Cai et al., 2010; Van Der Straaten et al., 2023). For instance, trade liberalization reduces price levels, boosts income, and effectively curtails consumption inequality (Cheong and Jung, 2021). Additionally, digital finance interventions were observed to reduce consumption inequality (de Moraes et al., 2023). Government policies also played a crucial role in shaping consumption inequality. Progressive consumption tax rates and personal income tax reforms were associated with reduced wealth and consumption relative deprivation(Khieu and Van Nguyen, 2020). Pro-poor transfers have also shown promise in mitigating rural-urban consumption inequality (Aaberge et al., 2019). Research on pension insurance revealed that income growth and income gap mitigation effects are mechanisms by which pension insurance reduces inter-individual relative deprivation, but the coexistence of multiple pension insurances also increased household consumption inequality(Li and Zang, 2022).

#### Economic impact of medical insurance

Studies have consistently shown that medical insurance can significantly influence consumption patterns and financial well-being. For example, Finkelstein et al. (2012) conducted a seminal study on the Oregon healthcare experiment, revealing that expanding health insurance coverage led to a substantial increase in healthcare utilization and a notable reduction in financial strain among low-income individuals. A study found the expansion of Medicaid coverage under the Affordable Care Act significantly increased the utilization of preventive health services and overall health status (Simon et al., 2017). The work of Cai et al. (2016) has shed light on the role of insurance in promoting household durable goods consumption in China, with urban households buying refrigerators, washing machines, and air conditioners, and rural households buying color TVs, refrigerators, washing machines, air conditioners, and computers. There is not much literature that focuses directly on the impact of health insurance on household consumption inequality. Johar et al. (2018) found that Indonesians’ access to health care was pro-rich in general but pro-poor at Puskesmas. Zhou and Huang (2021) compared the effectiveness of China’s basic health insurance in alleviating the relative deprivation of rural migrant workers and found that there are large differences in the fairness of the benefits of different health insurance policies. Unfortunately, their studies did not examine the effect of URMI.

#### The underlying mechanisms

Taken together, health service utilization and healthcare burden, precautionary savings, and insurance funding modalities may be the mechanisms through which URMI affects household consumption inequality. From the perspective of healthcare service utilization, Finkelstein et al. (2012) demonstrated that the Oregon healthcare reform program notably increased healthcare service utilization among low- and middle-income groups in the United States. However, Wagstaff et al. (2014) and Serrano-Lomelin et al.(2020) observed prevalent healthcare service utilization inequality resulting from health insurance expansion. With integrated health insurance, low-income groups may augment medical service utilization, potentially reducing instances of foregoing treatment(Hong et al., 2021). Yet, this could inadvertently escalate total medical costs. If the risk compensation mechanism fails to offset rising medical expenses, low-income groups may see a surge in out-of-pocket medical spending (Chang et al., 2021). Furthermore, rural-urban health insurance integration might exacerbate disparities in medical service utilization, as high-income groups with better health conditions potentially enjoy more comprehensive medical services, while the medical burden on low-income groups with poorer health conditions escalates (Fan et al., 2021; Jin et al., 2020). Analyzing precautionary savings, households with higher initial endowments may increase savings due to uncertainty surrounding future asset returns under the CRRA preference setting (Yin, 2021). URMI, offering superior treatment compared to the prior New Rural Cooperative Program, may prompt higher-income groups to allocate more precautionary savings, further amplifying inequality (Ma and Li, 2021). However, the asset erosion effect of premium payments may reduce disposable income for low-income households, dampening consumption (Fan et al., 2021). Lastly, the funding modalities transition from multi-tier to one-tier systems is a critical aspect of health insurance integration and will be gradually converted to the one-tier system as the income level of the household generally rises(Gao and Wang, 2017). The low-income group with limited ability to pay under the one-tier system may cause unbearable financial pressure due to the short-term substantial increase in premiums. By taking full account of the ability to pay off insured families, the multi-tier system can avoid unbearable financial pressure and reduce consumption inequality.

## 3. Research design

### 3.1 Data

The data utilized in this study originates from the China Family Panel Studies (CFPS), a longitudinal survey initiated by the China Social Science Survey Center at Peking University in 2010. Conducted biennially, CFPS captures a wide array of social, economic, demographic, educational, and health-related dynamics at the individual, family, and community levels, serving as a fundamental resource for numerous academic inquiries. For our analysis, we employ the recently released data spanning the years 2012, 2014, 2016, and 2018. Our data screening process involves several steps: First, we designate the “financial respondent” as the household head’s proxy and align household characteristics accordingly. Subsequently, we identify households that participated consistently across all four survey periods, ensuring data balance. Observations with urban domicile, health insurance enrolment status of not enrolled, missing control variables, and anomalies are excluded. Further refinement involves removing the highest and lowest 1 percent consumption levels samples and retaining households headed by individuals aged between 16 and 80 years. Following these procedures, our dataset comprises 17,092 observations from 4,273 households, providing a robust foundation for our analysis.

### 3.2 Variables

#### 3.2.1 Outcome variable

Household consumption inequality, denoted as *RD* (Relative Deprivation), is assessed using the Kakwani relative deprivation index, as outlined by Zhou and Huang(2021). Given the organizational framework of the current urban and rural health insurance system, which operates at the city or district level, households tend to select comparison samples from nearby geographical areas with higher consumption levels. Consequently, individuals residing in the same district as the surveyed household are chosen as the reference group for comparison. The consumption inequality index is then derived by contrasting the surveyed household with others in the district with higher consumption levels.

The calculation methodology is delineated as follows: Let *X* represent the reference group, with *N* denoting the total number of households within the group. These households are ranked based on ascending consumption levels, resulting in a consumption vector distribution represented as *C=(c_1_,c_2_,c_3_,…,c_N_)*, where *c_1_≤c_2_≤c_3_≤…≤ c_N_*. Here, *c_i_* represents the consumption level of the ith household within the group. By comparing the current consumption level of household *i* with that of other household *j* within the same cluster, the relative inequality can be expressed as:

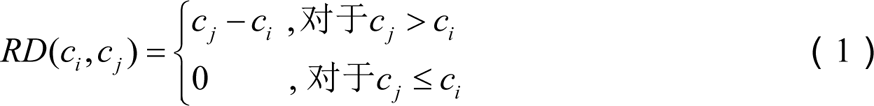

In equation (1), the indices *i* and *j* are constrained to satisfy 1 ≤ *i*, *j* ≤ *N*. This formulation signifies that a relative deprivation exists when the consumption level of household *j* exceeds that of household *i* (*c_j_*>*c_i_*). Conversely, no relative deprivation occurs when the consumption level of household j is equals or lower than that of household *i* (*c_j_* ≤ *c_i_*).

Let’s define some terms: *u_ci_^+^* represents the mean consumption across all households within the cohort that surpasses *c_i_*, *N_ci_^+^* signifies the total number of households consuming more than *c_i_*, and*γ_ci_^+^* represents the proportion of such households within the total sample. The Kakwani consumption relative deprivation index for household *i* is computed by summing *RD(c_i,_c_j_)* over all households *j* and dividing by the mean consumption of households within the cluster and the total sample size:

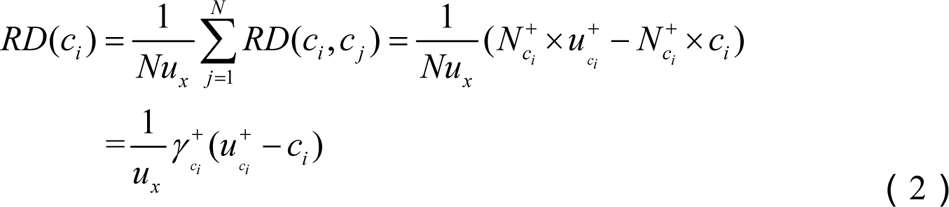

In equation (2), *RD*(*c_i_*) quantifies the relative deprivation experienced by household *i* concerning all other individuals within the same cohort who consume at a higher level than household *i*. This metric, bounded between 0 and 1, serves as a monotonically decreasing function of household consumption.

#### 3.2.2 Core explanatory variable

The implementation of the rural-urban health insurance coordination policy (*insurance*) serves as an exogenously driven government policy reform. This reform acts as a quasi-natural experiment, facilitating a comparative analysis of consumption inequality changes in rural areas pre- and post-policy implementation through a difference-in-differences model. The rural-urban health insurance integration policy is rolled out gradually across different regions. The determination of whether this policy is implemented in a specific region and the timing of implementation are established by gathering coordination implementation plans issued by respective government departments in each region, along with their issuance dates. The variable *insurance_ijt_* assigned a value of 1 for the current year and subsequent years if the policy has been enacted in the city where the household resides; otherwise, it is assigned a value of 0.

#### 3.2.3 Control variables

Drawing upon the research of Pak(2023) and Zhou et al.(2022), this paper selects control variables from two primary levels. Firstly, demographic characteristics encompass the gender of the household head (with males coded as 1 and females as 0), age and its square (to capture non-linear effects on consumption inequality), marital status (coded as 1 for married and 0 otherwise), and educational attainment (classified, with respective values assigned from 0 to 7). Secondly, household characteristics encompass indicators of income (logarithm of household income plus 1), assets (logarithm of financial assets and real estate value plus 1), population size, and participation in pension and health insurance schemes.

Descriptive statistics for each variable are presented in Table 1. According to the descriptive statistical results of the outcome variables, the treatment group exhibits higher levels of total consumption inequality, as well as inequality indices for subsistence, development, and enjoyment consumption, compared to the control group. It is suggested that the reform of the urban-rural health insurance integration policy may significantly contribute to the increased consumption inequality among rural residents. This hypothesis will be further tested and validated through subsequent empirical research. The descriptive statistics of the control variables reveal no significant disparities between the treatment and control groups across categories such as gender, age, marital status, income, financial assets, property, pension and health insurance participation,. These findings satisfy the balance test, indicating that the groups are well-matched in terms of these characteristics.

**Table 1.** Descriptive statistics of the variables.

|  | 2012 |  | 2014 |  | 2016 |  | 2018 |  |
| --- | --- | --- | --- | --- | --- | --- | --- | --- |
|  | t=0 | t=1 | t=0 | t=1 | t=0 | t=1 | t=0 | t=1 |
| RD | 0.402 | 0.433 | 0.413 | 0.501 | 0.422 | 0.465 | 0.379 | 0.455 |
| RDLI | 0.399 | 0.429 | 0.415 | 0.486 | 0.418 | 0.545 | 0.343 | 0.424 |
| RDDE | 0.559 | 0.568 | 0.518 | 0.481 | 0.532 | 0.541 | 0.536 | 0.550 |
| RDEN | 0.857 | 0.857 | 0.704 | 0.806 | 0.695 | 0.709 | 0.702 | 0.767 |
| gender | 0.423 | 0.586 | 0.522 | 0.544 | 0.514 | 0.536 | 0.517 | 0.586 |
| age | 48.10 | 48.31 | 50.61 | 51.43 | 51.46 | 51.97 | 52.80 | 53.31 |
| age2 | 24.89 | 25.24 | 27.31 | 28.24 | 28.38 | 28.40 | 29.88 | 30.18 |
| marr | 0.797 | 0.741 | 0.884 | 0.868 | 0.864 | 0.898 | 0.886 | 0.910 |
| edu | 1.525 | 1.345 | 1.765 | 1.886 | 1.563 | 1.268 | 2.027 | 1.787 |
| incom | 10.06 | 9.84 | 10.27 | 10.23 | 10.30 | 10.36 | 10.44 | 10.13 |
| financ | 0.375 | 0.325 | 0.512 | 0.505 | 0.584 | 0.423 | 0.647 | 0.614 |
| house | 1.314 | 1.225 | 2.355 | 2.224 | 2.487 | 2.594 | 3.003 | 2.478 |
| famnu | 3.995 | 4.224 | 3.857 | 3.798 | 3.793 | 3.975 | 3.607 | 3.884 |
| penins | 0.175 | 0.152 | 0.484 | 0.491 | 0.505 | 0.477 | 0.519 | 0.531 |
| medin | 0.899 | 0.879 | 0.927 | 0.904 | 0.933 | 0.905 | 0.937 | 0.955 |

Note: t=1 indicates the treatment group that implemented the rural-urban health insurance coordination, and t=0 indicates the control group that did not participate in the coordination.

### 3.3 Primary analysis of consumption inequality

Figure 1 presents an analysis of consumption relative deprivation trends in rural areas using data from CFPS from 2012 to 2018. Numerical examination reveals that household consumption inequality has remained within the range of 0.402 to 0.501, while the subsistence consumption inequality index, ranging from 0.399 to 0.545, closely mirrors the overall consumption inequality level. Notably, enjoyment consumption exhibits the highest level of inequality among various consumption types. A closer examination of the trend indicates an inverted U-shape distribution for both overall and subsistence consumption inequality indices, with peaks observed in 2014 and 2016, respectively, followed by gradual declines. This suggests a narrowing gap in consumption levels and subsistence expenditures among rural households in China in recent years. Conversely, development and enjoyment consumption inequality indices display a positive U-shaped trend, indicating a gradual expansion of such expenditures among rural households. Furthermore, households participating in the rural-urban health insurance coordination exhibit notably higher consumption relative deprivation compared to non-participating households, except for relative deprivation in development consumption.

**Figure 1.**
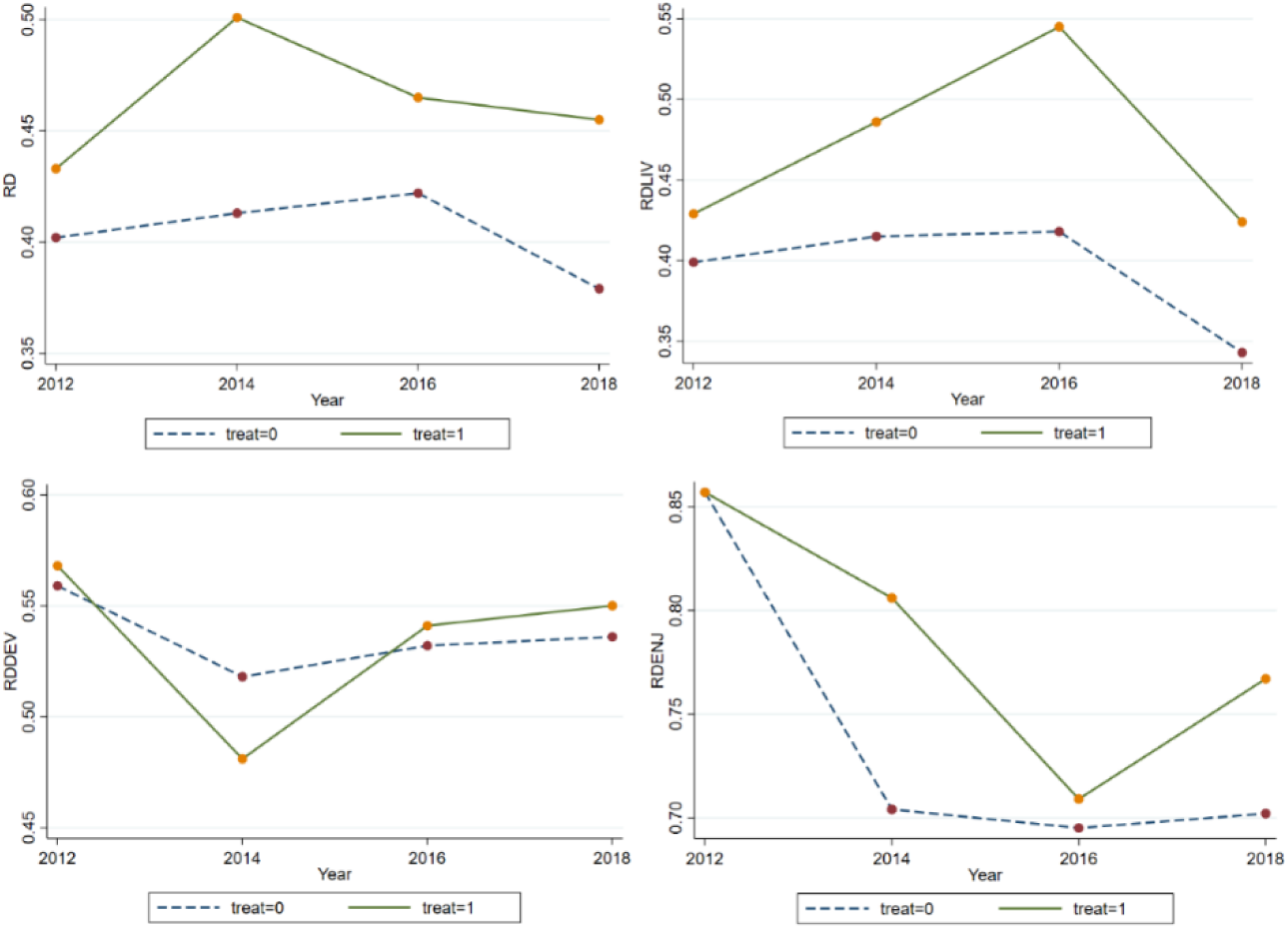
Overall and sub-consumption inequality indices. Notes: RD, RDLIV, RDDEV, and RDENJ represent the total consumption inequality index, the subsistence consumption inequality index, the development consumption inequality index, and the enjoyment consumption inequality index, respectively.

### 3.4 Empirical method

Given that the rural-urban health insurance coordination policy constitutes an exogenous shock for households, prior research has commonly employed the difference-in-differences-in-differences (DID) method to estimate changes in household health or healthcare burden (Chang et al., 2021; Ma and Li, 2021). However, owing to the policy’s gradual rollout across different regions, a staggered DID approach, as suggested by Beck et al. (2010), avoids explicitly defining a control group. Instead, it designates a control status based on whether rural-urban health insurance coordination is implemented in the district of each household before policy implementation. Subsequently, this control status transitions to an treatment group post-implementation. The model setup is delineated as follows:

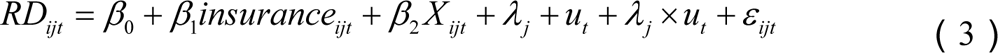

*RD_ijt_* denotes the degree of relative inequality in consumption (Kakwani relative deprivation index) in period *t* for the ith household located in district *j*; *insurance_ijt_* denotes whether rural-urban health insurance coordination has been implemented in period *t* in district *j* where household *i* is located and is assigned the value of 1 after the implementation of the policy, and 0 otherwise; and *X_ijt_* is the head and household characteristics variables, including gender, age and its square, marriage, education, household income, financial assets, property value, population size, pension insurance participation, health insurance participation. *λ_j_* and *μ_t_* are the area and time fixed effects, and the interaction terms of the area and the time fixed effects are also added to further rule out the effect of heterogeneous trends in consumption inequality across different areas. The coefficient *β_1_*measures the extent to which the implementation of the rural-urban health insurance coordination policy in rural areas affects relative consumption inequality.

## 4. Results

### 4.1 Baseline regression results

Table 2 presents the regression outcomes examining the impact of rural-urban health insurance coordination policy implementation on household consumption relative deprivation. Columns (1) to (3) progressively incorporate control for area and time fixed effects, along with household head and household characteristics. The results across the three columns consistently indicate a significantly positive effect of rural-urban health insurance integration implementation in rural areas on the Kakwani index at the 1% significance level. This finding suggests that the policy exacerbates consumption inequality among rural households.

**Table 2.**
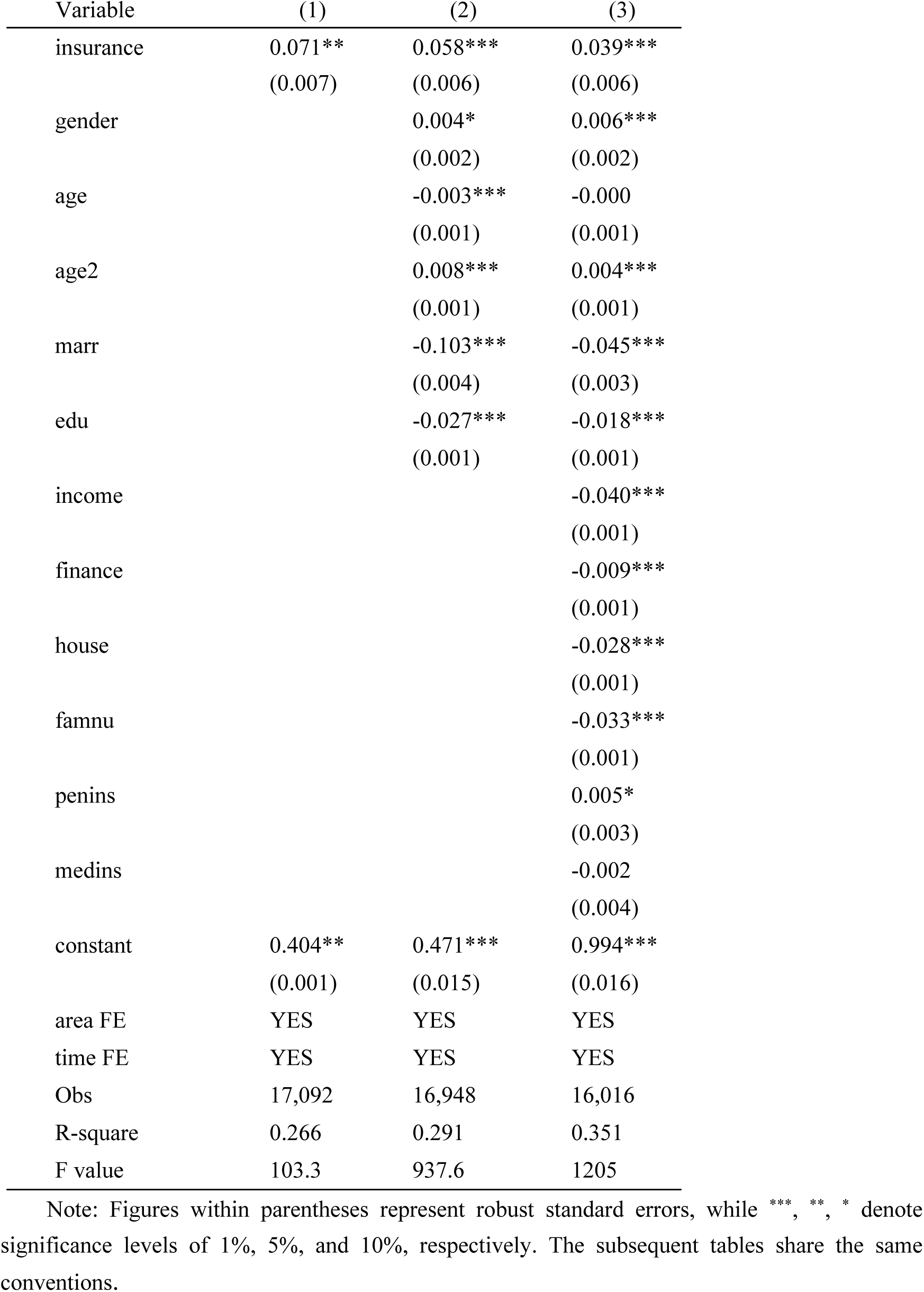
Benchmark regressions results.

About the characteristics of household head, the impact of age on rural household consumption relative deprivation shows a positive U-shaped trend, with consumption inequality being lower in middle age than in youth and old age, suggesting that the consumption level of middle-aged households is relatively high and that household consumption expenditures show heterogeneity over the life cycle. On average, consumption inequality is higher among male-headed households, a result that may be related to the generally lower willingness to consume among men. Married households have higher levels of household consumption expenditure because they are more risk-resistant and thus have less incentive to save. Educational attainment and income tend to be positively correlated, with higher education levels triggering higher levels of household consumption and thus lower levels of consumption inequality. In terms of household characteristics, population size has a significantly negative impact on consumption inequality, confirming that household size is an important means for rural households to protect themselves against economic risks. Income, financial assets, and property have a significantly negative impact on consumption inequality, as household income and assets increase, the wealth effect increases while liquidity constraints and precautionary saving incentives diminish, leading households to increase consumption. Participation in pension insurance in rural areas, on the other hand, significantly increases consumption inequality, consistent with the findings of Li and Zang (2022) that pension income in rural areas with relatively limited sources of income widens the gap in transfers between households and creates stronger perceptions of consumption deprivation among disadvantaged households that do not participate in pension insurance.

### 4.2 Regression tests by income quartile

To examine the consumption response across different income strata following the implementation of rural-urban integrated health insurance, household consumption expenditure’s logarithm serves as the outcome variable. Regression analysis is conducted both on the entire sample and on five sub-samples categorized by income quartiles: low-income, lower-middle-income, middle-income, middle-upper-income, and high-income groups based on the 20th, 40th, 60th, and 80th quartiles of household income. This approach aims to ascertain whether the integration of urban and rural health insurance disproportionately stimulates consumption among higher-income households. Table 3 showcases the results, where column (1) indicates a significant increase in rural residents’ consumption expenditures attributable to rural-urban health insurance coordination. Columns (2) to (6) further reveal the heterogeneous consumption effects across income levels: while health insurance coordination notably reduces consumption expenditures among low-income households, it significantly boosts consumption levels among middle-income, middle-upper-income, and high-income households. The magnitude of this effect escalates with higher household income quartiles, whereas the consumption expenditure of middle and low-income groups remains unaffected. This suggests that rural-urban health insurance coordination primarily unlocks consumption potential among rural middle- and high-income groups, with limited incentives for consumption expenditure among low-income groups.

**Table 3.** Regression results by income quartile.

| Variable | (1) | (2) | (3) | (4) | (5) | (6) |
| --- | --- | --- | --- | --- | --- | --- |
|  | Total | q0-q20 | q20-q40 | q40-q60 | q60-q80 | q80-q100 |
| insurance | 0.101**<br>(0.043) | -0.095*<br>(0.049) | -0.039<br>(0.043) | 0.112***<br>(0.038) | 0.173***<br>(0.036) | 0.221***<br>(0.039) |
| controls | YES | YES | YES | YES | YES | YES |
| area FE | YES | YES | YES | YES | YES | YES |
| time FE | YES | YES | YES | YES | YES | YES |
| Obs | 16,062 | 3,260 | 3,162 | 3,212 | 3,280 | 3,148 |
| R-square | 0.172 | 0.164 | 0.186 | 0.147 | 0.164 | 0.246 |
| F value | 40.15 | 37.29 | 40.23 | 25.24 | 22.83 | 49.45 |

### 4.3 Heterogeneity tests

The influence of rural-urban health insurance coordination on consumption inequality may encompass not only overall level effects but also heterogeneous structural effects across various consumption categories. Furthermore, its impact on the consumption gap among rural households may reveal nuanced age hierarchies. This section delves into the diverse effects of rural-urban health insurance coordination on rural consumption inequality.

1. Structural effects of consumption inequality. Consumption expenditure can be classified as subsistence, development, and enjoyment consumption. We use the Kakwani index, which measures the relative inequality indices of households’ subsistence, development, and enjoyment consumption respectively, as an outcome variable to examine the effects of the rural-urban health insurance coordination policy on relative deprivation in different types of consumption, and the regression results are shown in Table 4, columns (1) -column (3). It is easy to find that the rural-urban health insurance coordination policy has a significant expansion effect on both subsistence and enjoyment consumption inequality of rural households, and the comparison of the degree reveals that the expansion of subsistence consumption inequality is greater; on the contrary, development consumption inequality is not significantly affected by the coordination. The reasons for this may be: on the one hand, education expenditure is both the main item of development consumption in rural households and certainty for households with children in the education stage, and households generally invest less in development consumption other than that, thus the differences are limited. On the other hand, the consumption structure of rural households in China generally reflects a high proportion of subsistence consumption represented by food, and the implementation of rural-urban health insurance coordination in this context will have a greater impact on the release of consumption potential of the relatively high-income group, which has led to an intensification of the differentiation of subsistence consumption.
2. Life-cycle effect of consumption inequality. The impact of rural-urban health insurance coordination on consumption inequality across different age cohorts is examined in Table 4, columns (4)-(7). The analysis categorizes individuals into four age brackets: under 30 years old, 31-45 years old, 46-60 years old, and over 61 years old. Findings reveal a significant widening of consumption relative deprivation among individuals aged 46-60 and those aged 61 or older due to rural-urban health insurance coordination. Particularly pronounced is the consumption deprivation observed among households in the 61 or older age group, underscoring the considerable expansion of consumption inequality attributable to the implementation of health insurance coordination. This exacerbation of consumption inequality in the 46-60 or older age groups suggests a heightened sensitivity to policy changes among older individuals, likely influenced by their elevated health risks. Conversely, younger age groups exhibit comparatively lower sensitivity to alterations in health insurance policies owing to their reduced health risks.

**Table 4.** Heterogeneity tests results.

| Variable | (1) | (2) | (3) | (4) | (5) | (6) | (7) |
| --- | --- | --- | --- | --- | --- | --- | --- |
|  | RDLIV | RDDEV | RDENJ | under 30 | 31-45 | 46-60 | over 61 |
| insurance | 0.037***<br>(0.006) | -0.003<br>(0.007) | 0.024***<br>(0.007) | 0.018<br>(0.022) | 0.016<br>(0.011) | 0.033***<br>(0.009) | 0.058***<br>(0.011) |
| controls | YES | YES | YES | YES | YES | YES | YES |
| area FE | YES | YES | YES | YES | YES | YES | YES |
| time FE | YES | YES | YES | YES | YES | YES | YES |
| Obs | 16,016 | 16,016 | 16,016 | 1,676 | 4,956 | 6,824 | 4,208 |
| R-square | 0.330 | 0.311 | 0.288 | 0.275 | 0.257 | 0.303 | 0.407 |
| F value | 68.02 | 578.71 | 612.10 | 27.31 | 137.40 | 339.20 | 303.50 |

### 4.4 Robustness tests

#### 4.4.1 Parallel trend test

If there is no discernible disparity in consumption inequality trends between the treatment and control groups before the enactment of the rural-urban health insurance coordination policy, yet a significant discrepancy emerges post-implementation, it suggests that the policy itself influences consumption inequality. Given variations in the timing of policy implementation across regions, this study employs the event study method to conduct a multi-period difference-in-differences (DID) analysis. This involves introducing a dummy variable spanning from four periods before to two periods after policy implementation, alongside the interaction term of the rural-urban health insurance coordination policy, to regress on consumption inequality. The model is formulated as follows:

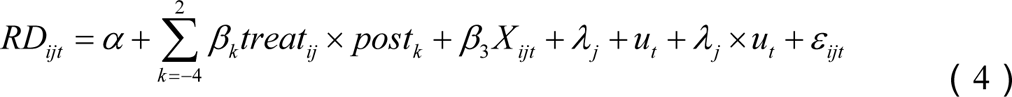

In equation (4), *treat_ij_*denotes whether household *i* located in district *j* has implemented rural-urban health insurance coordination during the survey period, with a value of 1 assigned to implementation and 0 otherwise. The variable *post_k_* represents the year dummy variable, ranging from -4 to 2, indicating 1-4 years before policy implementation, the implementation year, and 1-2 years post-implementation, respectively. Other parameters are defined analogously to equation (3). Parameter *β_k_* reflects the impact of insurance coordination implementation on consumption disparities between coordinated and non-coordinated regions, with the 4th year pre-implementation serving as the base period. As depicted in Figure 2, before coordination, the 95% confidence intervals of estimated coefficients *β_-3_*, *β_-2_* and *β_-1_* do not significantly deviate from 0, indicating satisfactory parallel trends between treatment and control groups. Subsequently, following coordination, the 95% confidence intervals of *β_1_* and *β_2_*gradually and significantly diverge from 0, supporting the assertion that rural-urban health insurance coordination exacerbates consumption relative deprivation in rural areas as indicated in the benchmark regression.

**Figure 2.**
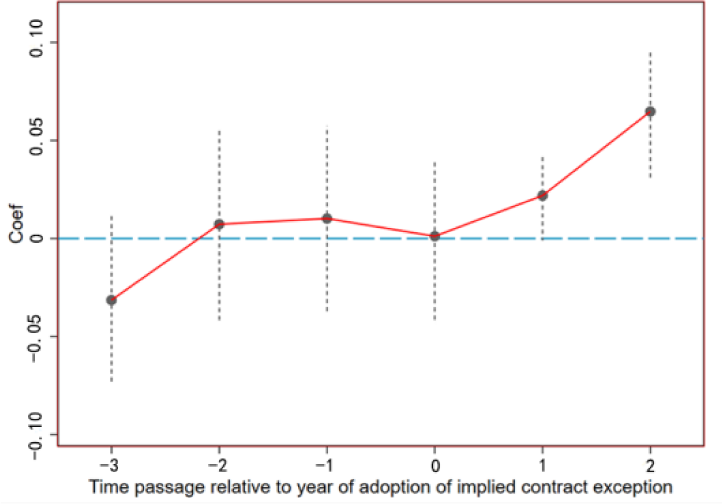
Parallel trend test.

#### 4.4.2 Placebo test

In this study, we utilize a non-parametric permutation test to mitigate the influence of other policy shocks. Additionally, we conduct a placebo test by randomly assigning households in the sample to either implement or not implement the rural-urban health insurance coordination policy. The sample comprises 1,388 households enrolled in the rural-urban health insurance scheme. Among these, 1,388 households are randomly designated as the false treatment group, while the remaining households constitute the false control group. This process is repeated 500 times to generate 500 randomized treatment and control groups, which are subsequently re-estimated using the generated randomized samples following equation (3). Given that the false treatment group is not grounded in actual implementation, health insurance integration theoretically exerts no significant impact on consumption inequality (*β_1_^false^*=0). Conversely, a statistically significant deviation of *β_1_^false^* from 0 indicates potential identification bias in the baseline model. Test results illustrated in Figure 3 demonstrate that the mean values of the estimated coefficients derived from the 500 regressions predominantly cluster around 0. Conversely, the estimated coefficients from the benchmark regression (depicted by the red dotted line) distinctly deviate from this pattern, underscoring the significant causal effect of the rural-urban health insurance integration policy on rural consumption relative deprivation.

**Figure 3.**
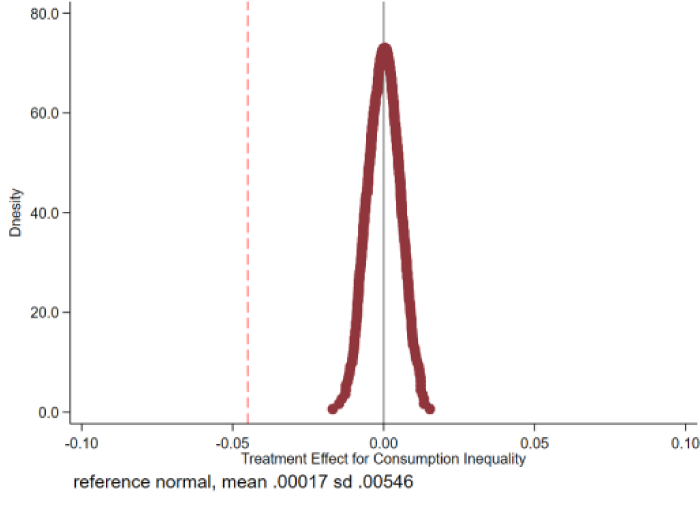
Non-parametric permutation test.

#### 4.4.3 PSM-DID test

The DID model addresses endogeneity issues stemming from omitted unobservable characteristics, while the Propensity Score Matching (PSM) approach corrects selectivity bias arising from observable variables. Initially, household head and household-level matching variables were employed to filter samples within the treatment and control groups using propensity score matching via 1:1 nearest neighbor matching, kernel matching, and radius matching techniques. The matched samples were then subjected to staggered DID estimation. Results presented in Table 5, columns (1)-(3), underscore the significant positive impact of implementing an integrated rural-urban health insurance policy in rural areas on consumption inequality.

**Table 5.** Robustness tests.

| Variable | PSM-DID test |  |  | Tobit-DID model | Substitute outcome variable |
| --- | --- | --- | --- | --- | --- |
|  | Nearest neighbor matching | Kernel matching | Radius matching |  |  |
|  | (1) | (2) | (3) | ( 4 ) | ( 5 ) |
| insurance | 0.037***<br>(0.006) | 0.031***<br>(0.007) | 0.024***<br>(0.007) | 0.067***<br>(0.011) | 0.055**<br>(0.023) |
| controls | YES | YES | YES | YES | YES |
| area FE | YES | YES | YES | YES | YES |
| time FE | YES | YES | YES | YES | YES |
| Obs | 11,104 | 11,104 | 11,104 | 16,016 | 16,016 |
| R-square | 0.312 | 0.291 | 0.324 | 0.236 | 0.235 |
| F value | 68.02 | 578.7 | 612.1 | 68.02 | 68.02 |

#### 4.4.4 Substitution identification method or outcome variable

Firstly, given that the outcome variable ranges between 0 and 1, this study employs the staggered DID method based on the restricted outcome variable (Tobit) model to reexamine the relationship between the implementation of rural-urban health insurance coordination policy and consumption inequality in rural areas. Column (4) of Table 5 reveals a significantly positive coefficient. Secondly, refer to Deng and Yang (2019), the Kakwani relative deprivation index is transformed into five ordered levels of consumption deprivation by dividing the index into intervals of 0.2. This transformed index is then re-evaluated as an outcome variable utilizing the Ordered Probit model. Column (5) of Table 5 demonstrates a significantly positive coefficient, consistent with the findings of the baseline regression analysis.

## 5. Mechanism analysis

This study delves into the potential pathways by which rural-urban health insurance coordination influences consumption inequality. It examines the impact on health service utilization and healthcare burdens, precautionary savings, and variations in health insurance funding modalities.

### 5.1 Healthcare service utilization

This section investigates the impact of rural-urban health insurance coordination on healthcare motivation and service utilization across various income strata. Drawing insights from Chang et al. (2021), who utilize outpatient visit probability and hospitalization probability to gauge healthcare motivation, as well as the logarithm of medical out-of-pocket expenses and the ratio of medical out-of-pocket expenses to household non-medical consumption to measure healthcare service utilization, regression analysis is conducted by introducing the interaction term of rural-urban health insurance coordination with logarithmic household income. Results presented in Table 6 reveal noteworthy patterns. In columns (1) and (2) analyzing healthcare motivation, the implementation of health insurance coordination significantly boosts the likelihood of household consultations and hospitalizations. However, this enhancement effect diminishes significantly with rising income levels, suggesting that while rural-urban health insurance coordination bolsters healthcare-seeking among rural households, its impact is more pronounced for low-income groups. From the analysis of health service utilization in columns (3) and (4), it can be seen that after the implementation of coordination, out-of-pocket expenses and the ratio of medical out-of-pocket expenses to household non-medical consumption both increased at the 1% test level. The proportion of out-of-pocket expenses also showed a significant decrease with increasing income, indicating that the integration correspondingly increased the medical burden on low-income groups. This phenomenon can be attributed to the fact that low-income individuals, constrained by financial limitations, often adopt a medical avoidance mindset. The integration of health insurance substantially increases healthcare accessibility for these groups, thereby unleashing pent-up medical needs previously suppressed by budget constraints. Consequently, there is a surge in medical out-of-pocket expenses, leading to a crowding-out effect on current household consumption. Conversely, the rise in out-of-pocket medical expenses for high-income households remains more moderate, indicating that increased medical service utilization among this cohort has minimal crowding-out effects on household consumption.

**Table 6.** Impact of the rural-urban health insurance integration policy on health service utilization.

| Variable | (1) | (2) | (3) | (4) |
| --- | --- | --- | --- | --- |
|  | healthcare motivation<br>outpatient | hospitaliz | health service utilization<br>out-of- | ratio of out-of- |
| insurance | 0.118**<br>(0.052) | 0.115**<br>(0.055) | 0.109***<br>(0.027) | 0.146***<br>(0.031) |
| insurance×lninco | -0.008**<br>(0.004) | -0.007**<br>(0.004) | -0.013<br>(0.009) | -0.011***<br>(0.003) |
| controls | YES | YES | YES | YES |
| area FE | YES | YES | YES | YES |
| time FE | YES | YES | YES | YES |
| Obs | 15,033 | 14,509 | 13,407 | 13,407 |
| R-square | 0.333 | 0.324 | 0.335 | 0.316 |
| F value | 18.42 | 22.72 | 67.58 | 112.30 |

Table 7 presents the dynamic effects of policy implementation on healthcare motivations and service utilization. In terms of healthcare incentives, columns (1) and (2) indicate that before the implementation of the coordinated health insurance scheme, the probability of outpatient and hospitalization is significantly higher for high-income households compared to low-income households during periods 1-2 and 2-3. However, no statistically significant differences exist in consultation and hospitalization probabilities between income groups during the implementation period of the coordinated scheme. Furthermore, household consultation and hospitalization probabilities decrease with rising income during the first year of policy implementation, with policy effects no longer significant after the second period of coordination. This suggests that the short-term expansionary impact of rural-urban health insurance coordination on healthcare accessibility for low-income households is notably pronounced. Regarding healthcare service utilization, columns (3) and (4) demonstrate that before healthcare insurance coordination, both out-of-pocket expenses and the proportion of out-of-pocket expenses increased significantly with household income, with healthcare consumption notably higher among high-income groups. However, in the year of policy implementation and beyond, out-of-pocket expenses no longer exhibit significant statistical differences across income groups, and the proportion of out-of-pocket expenses gradually decreases with income. This indicates that the rural-urban health insurance coordination policy has mitigated disparities in absolute out-of-pocket medical expenses between income groups. Nevertheless, the burden of out-of-pocket medical expenses on low-income households remains heavier than on high-income households, exacerbating inequality by squeezing disposable income among low-income groups and suppressing consumption expenditures to a greater extent.

**Table 7.** Dynamic impact of rural-urban health insurance integration policy on health service utilization.

| Variable | (1) | (2) | (3) | (4) |
| --- | --- | --- | --- | --- |
|  | healthcare motivation |  | health service utilization |  |
|  | outpatient | hospitaliza | out-of- | ratio of out-of- |
| post-4×lnincome | 0.000<br>(0.001) | -0.001<br>(0.001) | 0.008***<br>(0.001) | 0.003***<br>(0.001) |
| post-3×lnincome | 0.001<br>(0.001) | 0.003***<br>(0.001) | 0.005**<br>(0.002) | 0.003***<br>(0.001) |
| post-2×lnincome | 0.003***<br>(0.001) | 0.002**<br>(0.001) | 0.011*<br>(0.006) | 0.003***<br>(0.001) |
| post-1×lnincome | 0.002**<br>(0.001) | 0.000<br>(0.001) | 0.015**<br>(0.006) | 0.004***<br>(0.001) |
| post0×lnincome | -0.007<br>(0.005) | 0.001<br>(0.004) | -0.017<br>(0.035) | -0.013**<br>(0.006) |
| post1×lnincome | -0.013***<br>(0.005) | -0.003***<br>(0.001) | -0.033<br>(0.037) | -0.016***<br>(0.006) |
| post2×lnincome | -0.007<br>(0.006) | 0.001<br>(0.005) | -0.017<br>(0.045) | -0.015*<br>(0.008) |
| post3×lnincome | -0.002<br>(0.006) | 0.007<br>(0.005) | 0.002<br>(0.045) | -0.017*<br>(0.009) |
| controls | YES | YES | YES | YES |
| area FE | YES | YES | YES | YES |
| time FE | YES | YES | YES | YES |
| Obs | 15,033 | 14,509 | 13,090 | 13,038 |
| R-square | 0.110 | 0.112 | 0.219 | 0.212 |
| F value | 19.69 | 19.55 | 58.12 | 53.69 |

### 5.2 Precautionary savings

Households with higher initial endowments tend to engage in more precautionary savings (Yin, 2021). Does rural-urban coordinated health insurance have a more pronounced effect on releasing precautionary savings in the high-consumption group compared to the low- and middle-consumption groups? Sample grouping regressions are conducted with precautionary savings as the outcome variable, categorized by consumption level quintiles. Precautionary saving is proxied by the proportion of liquid assets held by households. The regression outcomes in Table 8 reveal that for consumption level quintiles 0-20 and 20-40, the regression coefficients of rural-urban coordinated health insurance on precautionary savings are statistically insignificant. However, for quintiles 40-60, 60-80, and 80-100, rural-urban coordinated health insurance significantly reduces household precautionary savings, with this releasing effect expanding as consumption levels increase. This underscores the higher prevalence of precautionary savings among middle- and high-consumption groups in rural areas, and the subsequent release of savings motivation following the implementation of health insurance integration due to enhanced health insurance benefits, thereby promoting consumption. Conversely, it is challenging to diminish the incentive for precautionary savings among low-income groups, and the integration of the health insurance scheme has yet to unlock their consumption potential.

**Table 8.** Impact of the rural-urban health insurance integration policy on precautionary savings.

| Variable | (1) | (2) | (3) | (4) | (5) |
| --- | --- | --- | --- | --- | --- |
|  | Precautionary savings |  |  |  |  |
|  | q0-q20 | q20-q40 | q40-q60 | q60-q80 | q80-q100 |
| insurance | -0.001<br>(0.004) | -0.005<br>(0.005) | -0.010**<br>(0.005) | -0.012**<br>(0.005) | -0.015***<br>(0.005) |
| controls | YES | YES | YES | YES | YES |
| area FE | YES | YES | YES | YES | YES |
| time E | YES | YES | YES | YES | YES |
| Obs | 3,991 | 3,039 | 3,062 | 3,154 | 3,189 |
| R-square | 0.990 | 0.979 | 0.972 | 0.956 | 0.961 |
| F value | 93.97 | 153.3 | 326.8 | 1260 | 4642 |

### 5.3. Funding modalities analysis

Two primary funding modalities exist for rural-urban integrated health insurance: the one-tier system and the multi-tier system. The one-tier system applies the principle that all participants pay the same premiums and enjoy the same treatment, while the multi-tier system allows adult participants to choose between two or more tiers of contributions on their own and follows the principle of linking benefits to contributions. This section examines whether these different financing modes influence the degree of consumption inequality. The sample is divided into one-tier and multi-tier regions, and regression analysis is conducted according to equation (3). The results presented in Table 9 reveal that rural-urban health insurance coordination has a significantly higher impact on consumption inequality in one-tier regions compared to multi-tier regions, suggesting that the one-tier model exacerbates consumption inequality to a greater extent in rural areas. A plausible explanation is that, amidst low disposable income, the multi-tiered model accounts for income disparities among rural households by offering flexible contribution standards, enabling low-income households to choose affordable participation options based on their financial circumstances. This approach alleviates budget constraints, prevents premium increases from crowding out non-medical consumption, and empowers individuals with the autonomy to make independent choices. Consequently, this contributes to reducing consumption inequality between urban and rural areas. Moreover, individual autonomy in selecting contribution standards helps mitigate moral hazards and excessive medical demands resulting from passive payment of high premiums, fostering moderate consumption growth while maintaining stability. This underscores the effectiveness of the multi-tiered system as a transitional model for healthcare integration within the rural-urban healthcare framework, tailored to local needs.

**Table 9.** Regression results distinguishing between funding modalities.

|  | (1) | (2) |
| --- | --- | --- |
|  | One-tier | Multi-tier |
| insurance | 0.041***<br>(0.007) | 0.036***<br>(0.013) |
| controls | YES | YES |
| area FE | YES | YES |
| time FE | YES | YES |
| Obs | 12,842 | 2,108 |
| R-square | 0.318 | 0.363 |
| F value | 1262 | 112.6 |

## 6. Discussion

Focusing on the impact of the rural-urban health insurance coordination policy on rural households’ consumption relative deprivation and its mechanism, this paper uses a staggered difference-in-differences model based on data from the China Family Panel Studies (CFPS) for the four periods from 2012 to 2018. Results reveal a significant increase in household consumption inequality due to the policy. While the integration of rural-urban health insurance aligns contribution amounts, compensation standards, and health insurance treatment, potentially enhancing equality of opportunity in health insurance access for rural participants, it does not necessarily translate into more equitable consumption outcomes. These findings are consistent with the results of Fan et al. (2021), Chang et al. (2021), and Jin et al. (2020), who found there is a reverse subsidy mechanism in the integration of urban and rural health insurance, which is reflected in the fact that high-income groups enjoy more adequate healthcare services while the healthcare burden of low-income groups has increased significantly. This paper differs from them in that it examines for the first time the inequality effects of health insurance integration from a consumer welfare perspective, whereas the aforementioned articles are based on health inequality and healthcare utilization inequality perspectives. Regression tests by income quartile indicate that this discrepancy stems from the integrated health insurance disproportionately benefiting higher-income rural groups in terms of consumption level augmentation compared to lower-income groups, which is a departure from the original policy intent of alleviating the burden on vulnerable groups. Robustness tests, including parallel trend tests, non-parametric permutation tests, PSM-DID analyses, and variations in identification methods and outcome variables, confirm the consistency of results. To mitigate relative deprivation among rural low-income groups and foster overall well-being, several measures should be proposed to enhance the fairness of the rural-urban integrated health insurance system, with heightened attention to ensuring equitable benefits for the low-income groups.

Analysis of consumption structure reveals a notable expansion effect of urban and rural health insurance integration on subsistence and enjoyment consumption inequality among rural households, with minimal impact on development consumption inequality. The unleashed consumption potential from health insurance integration primarily stimulates growth in subsistence consumption among higher-income groups. This implies that it is difficult to achieve an increase in the consumption level and structural upgrading of rural residents by relying only on health insurance coordination (Jin et al., 2020) and that it is necessary to give rural areas more complementary consumption support policies to enhance consumption capacity and promote parity in the growth of high-quality consumption. Previous studies showed that household social insurance participation status and consumption deprivation are often related to the stage of the household life cycle in which they live (Li and Zang, 2022; Deng and Yang, 2019). This study highlights that health insurance integration primarily exacerbates consumption disparities among middle-aged and elderly groups, underscoring the need for fair distribution of benefits, particularly among the elderly and disadvantaged groups. This exacerbation of consumption inequality among individuals aged 46-60 and those aged over 61 suggests a heightened sensitivity to policy changes among older individuals, likely influenced by their elevated health risks. Conversely, younger age groups exhibit comparatively lower sensitivity to alterations in health insurance policies owing to their reduced health risks. This assertion is supported by studies, which indicate that older age cohorts are more responsive to psychological shifts in health insurance policies, leading to more pronounced consumption responses (Hong et al., 2021). It is imperative to acknowledge the vulnerability of older age cohorts to health-related challenges, poverty, and general susceptibility. Consequently, efforts aimed at enhancing health insurance integration should prioritize equitable distribution of benefits among these vulnerable groups, especially the elderly.

Channel analysis reveals a pro-poor trend in health service utilization resulting from rural-urban health insurance integration, facilitating low-income groups in fulfilling long-suppressed medical consumption needs and increasing the proportion of out-of-pocket medical care. This is consistent with the findings of Finkelstein (2011), which found that the healthcare reform in the USA significantly improved healthcare utilization in the low and middle-income brackets. Considering precautionary savings, we find that health insurance integration predominantly releases precautionary savings among middle- and high-consumption level households rather than among the poor. This suggests that households with higher initial endowments tend to engage in more precautionary savings (Yin, 2021) and health insurance consolidation beneficially frees up precautionary savings in this group. In addition, many studies have considered the rationalization of funding modalities settings, but the conclusions have not been consistent (Gao and Wang, 2017; Fan et al., 2021). This research supposes that in the context of generally low disposable incomes among rural residents, a multi-tiered system provides low-income households with the convenience of choosing an affordable contribution rate based on their financial situation. Conversely, the one-tier model exacerbates rural consumption inequality by intensifying financial constraints among poor households. To effectively alleviate the sense of consumption deprivation among low-income groups, it is necessary to take measures to address the long-standing demand for healthcare by releasing it in an orderly manner, alleviating the incentive for precautionary savings, and improving the income redistributive function of the health insurance system.

## 7. Conclusions

This study investigates the impact of the rural-urban health insurance coordination policy on consumption inequality among rural households and its underlying mechanisms using data from the China Family Panel Studies from 2012 to 2018, employing a staggered difference-in-differences model. The findings indicate a significant increase in household consumption relative deprivation due to the policy while ensuring equitable access to enhanced health insurance benefits for rural households. Health insurance coordination not only elevates consumption levels among middle- and high-income groups but also reduces consumption expenditures for low-income groups. The conclusions drawn from this paper reveal that despite the objectives of the rural-urban health insurance integration to dissolve household registration barriers and promote equity in healthcare access, it has yet to effectively narrow the consumption gap in rural areas, which should be a matter of concern for policymakers.

## Data Availability

The datasets for this study can be found in the China Family Panel Studies (CFPS) database (isss.pku.edu.cn/cfps/). The CFPS project was conducted by the China Social Science Survey Center of Peking University and publicly available.

## Ethical approval

This article does not contain any studies with human participants performed by any of the authors.

